# Resting EEG spectral slopes are associated with age-related differences in information processing speed

**DOI:** 10.1101/2021.02.12.21251655

**Authors:** A. Pathania, M.J. Euler, M. Clark, R. Cowan, K. Duff, K.R. Lohse

**Affiliations:** Department of Health and Kinesiology, University of Utah; Department of Psychology, University of Utah; Department of Neurology, University of Utah; Program in Physical Therapy and Department of Neurology, Washington University School of Medicine in Saint Louis

## Abstract

**Background:** Previous research has shown the slope of the EEG power spectrum differentiates between older and younger adults in various experimental cognitive tasks. Here, we extend that work, assessing the relation between the EEG power spectrum and performance on the Repeatable Battery for the Assessment of Neuropsychological Status (RBANS), a widely-used neuropsychological instrument that assesses a broad range of cognitive domains.

**Methods:** Forty-four participants (21 younger adults, 23 older adults) completed the RBANS with EEG data collected at-rest. Using spectral parameterization, we tested the mediating effect of the spectral slope on age-related differences in subsequent cognitive task performance.

**Results:** Older adults performed reliably worse on the RBANS overall, and on the Attention and Delayed Memory domains. However, evidence of mediation was only found for the Coding subtest, a measure of information processing speed.

**Conclusions:** We found some evidence that the slope of the resting EEG power spectrum mediated age-related differences in cognition. These effects were evident only in tasks requiring speeded processing, whereas this effect was not statistically significant for delayed memory, even though age-related differences were present.

---

Aging is broadly associated with declines in cognitive and motor abilities. Although the average pattern is for a decline in cognitive function with age, these changes are heterogeneous, such that within “normal” aging, there are substantial individual differences in both average performance and changes in performance over time (Habib et al., 2007). Some individuals experience declines so severe that they exceed the range of healthy aging, leading to mild cognitive impairment (MCI) or even dementia (Tucker-Drob, 2019). These declines can be attributed to numerous neurophysiological changes over the lifespan (Bäckman et al., 2006; Cabeza et al., 2016; Hedden & Gabrieli, 2004). Furthermore, these physical changes are broad and influence many different psychological processes, such as executive functions (Buckner, 2004), affective regulation (Fiske et al., 2009), and memory formation (Nyberg et al., 2012).

Although numerous studies have documented the wide range of individual differences in older adults’ cognitive function (Andrews et al., 2002; Dekhtyar et al., 2017; Lindenberger, 2014), the causes of these declines are unclear. Indeed, it is likely that age-related declines are multi-causal (Harada et al., 2013), further complicating the issue. Across different domains of cognition and changes in anatomical structures, one unifying way of understanding this problem from a functional perspective is the *neural noise hypothesis*. The neural noise hypothesis suggests that many (not all) age-related changes in cognition might be explained by a reduction in the signal-to-noise ratio of neural circuits with increased chronological age (Cremer & Zeef, 1987; Voytek & Knight, 2015; Waschke et al., 2017). Recently, researchers have proposed that the slope of the EEG power spectrum may be a viable correlate of neural noise.

## Characterizing the EEG Power Spectrum

The slope of EEG power spectrum exhibits a pattern of 1/*f* noise, which is a common finding in many biological, physical, and even social systems (He, 2014; Ward & Greenwood, 2007). We can think of 1/*f* noise as a particular kind of pattern where low frequencies have the most power in the signal, and power decreases at higher frequencies; that is, there is a negative relationship between power and frequency, as is common in human EEG (Cohen, 2014), which follows the form:

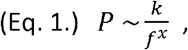

where the power in a signal (*P*) is inversely proportional to the frequency (1/*f*), with this proportionality determined by a decay parameter (*x*), hereafter referred to as the exponent, and a constant (*k*), hereafter referred to as the offset. When transformed into log-log space, this relationship becomes a linear function:

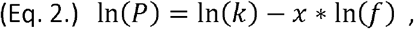

such that power in the signal monotonically decreases as the frequency increases, as shown in Figure 1. That is, in the resulting linear function, the exponent is now the slope of the line relating power to frequency. Many systems show this proportionality (with some relabeling of terms) from geology (Holliger, 1996), to music (Voss & Clarke, 1975), to human cognition (Gilden et al., 1995; Wagenmakers et al., 2004).

**Figure 1.**
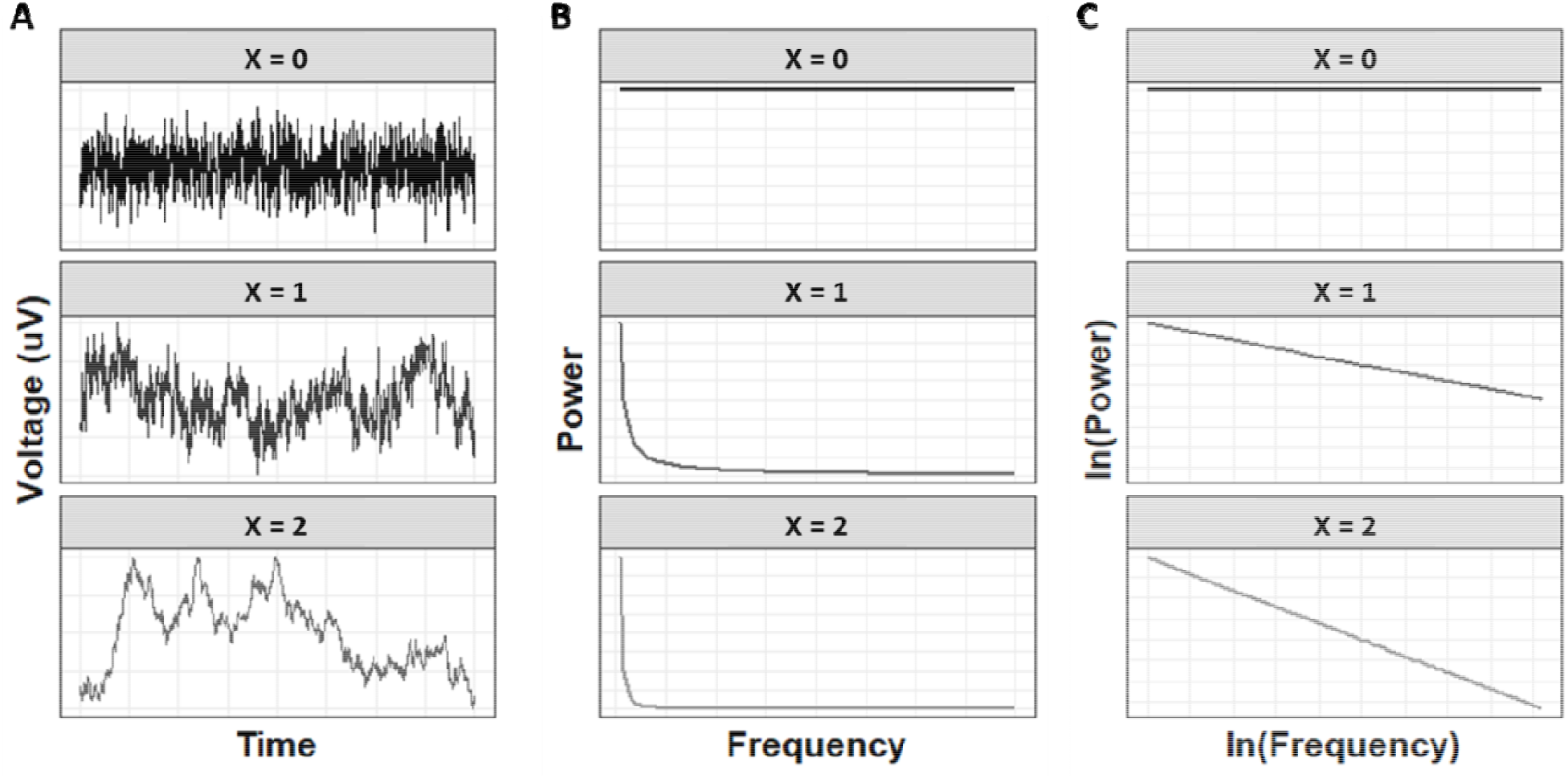
Three different time-series of hypothetical electrophysiological data. (A) Voltages are shown over time. (B) Following a fast Fourier transform, an exponential function (—) is fit to the raw power spectrum. (C) By log-transforming the Power and the Frequency of the power spectrum, we can now approximate this relationship with a linear function (). These transformations are shown for three different cases where the value of the exponent changes from 0 (shown in black, representing “white” noise), to 1 (shown in dark grey, representing “pink” noise), to 2 (shown in light grey, representing Brownian or “red” noise).

In the human EEG, a large proportion of the signal follows a broad-band 1/*f* pattern, with deviations away from this pattern reflecting periodic activity in the canonical frequency bands (e.g., alpha, beta, delta). Whereas decades of research has demonstrated relations between various oscillatory processes and specific psychological states or correlates, the psychological significance of the aperiodic (1/*f*) component of the power spectrum has only recently become clear. Because the 1/*f* pattern is so prevalent across different types of systems, it was often regarded as a form of background noise, with limited functional significance (see He, 2014 for a review). However, in the last several decades, key studies have established a plausible physiological basis for the spectral slope (Freeman & Zhai, 2009; He et al., 2010) and a growing number of functional correlates (Ouyang et al., 2020; Voytek et al., 2015).

Most recently, studies have provided evidence that the spectral slope reflects the balance of excitation and inhibition (E:I) in the brain, such that flatter slopes indicate an increase in random excitatory spiking, a potential source of noise in the signal (Gao et al., 2017). Although those findings, and much of this literature, has involved invasive recordings from local field potentials or electrocorticography, more recent research has extended this to scalp EEG, demonstrating that anesthetic agents with known (and opposite) influences on excitation and inhibition modulate the spectral slope in the expected direction (Waschke et al., 2021). Other aspects of the electrophysiological power spectrum are increasingly understood as well, such as the relation of the spectral offset to neuronal firing rates (Manning et al., 2009), and the relation of the characteristic bend in the slope (the “knee” frequency) to the timescale of neural dynamics (Gao et al., 2020). This empirical work aligns with simulation studies, which have shown that the slope and other spectral features can be well-modelled as a function of local neuronal spikes and aggregate synaptic dynamics (Freeman & Zhai, 2009; Gao et al., 2020; Miller et al., 2009). Altogether, recent basic science on the spectral slope provides a compelling basis for considering it a viable index of neural noise (Tran et al., 2020).

## Functional and Age-Related Correlates of the Spectral Slope

Simultaneous with these developments in understanding the mechanistic basis of the spectral slope, many studies have identified important functional correlates. For instance, the spectral slope has been observed to change across the lifespan (Dave et al., 2018; Donoghue et al., 2020; Schaworonkow & Voytek, 2021; Tran et al., 2020; Voytek & Knight, 2015), across a variety of tasks (Ouyang et al., 2020; Pathania et al., 2021; Voytek et al., 2015), across state-dependent variations in sleep (Freeman & Zhai, 2009; Leemburg et al., 2018; Miskovic et al., 2019), and during anesthesia (Gao et al., 2017; Waschke et al., 2021). Furthermore, recent evidence points to the EEG spectral slope being a product of both fast networks representing the local dynamics and slow networks representing the more distributed recurrent connections. These two types of networks balance the excitation and inhibition to maximize the information processing capabilities (Chaudhuri et al., 2018). Thus, the 1/*f* component of the power spectrum is not only a slowly changing tonic signal, but an important physiological signal that can change quite quickly given the demands of a task and the state of an individual.

With respect to aging specifically, several recent studies have shown flattening of the spectral slope associated with differences in cognitive performance, or other aspects of brain functioning (Dave et al., 2018; McNair et al., 2019; Tran et al., 2020; Waschke et al., 2017). Voytek and colleagues (2015) conducted one of the first studies in this area, demonstrating that differences in the EEG spectral slope at rest were not only related to age differences in working memory performance, but the slope specifically *mediated* that effect. That is, older adults did not merely differ in both performance and slope values from younger adults, but the behavioral differences could be accounted for statistically by the slope effect alone. In the time since, many studies have replicated and expanded upon this basic effect. For example, in the neural domain, flatter slopes in older adults have been related to reductions in the amplitude of the N400 event-related potential (ERP) during language processing (Dave et al., 2018), delayed auditory P2 ERP latencies during tone discrimination (McNair et al., 2019), and changes in various measures of single-trial neural variability (Tran et al., 2020; Waschke et al., 2017). Behaviorally, some of these and other studies have linked flatter slopes in older adults to poorer performances on tasks of spatial attention (Tran et al., 2020) and visual-short-term memory (Thuwal et al., 2021). Still other work has linked changes in the spectral offset to poorer cognitive control across the adult age range (Clements et al., 2021), while spectral slope has been linked to speeded target detection in adults aged 40 and below (Ouyang et al., 2020).

Overall, these studies demonstrate a consistent pattern of flatter slopes in older adults, which are linked to changes in functioning across a variety of tasks, and in line with the neural noise hypothesis. Notwithstanding this progress, there are several outstanding questions, which if addressed, could further enhance understanding of the functional significance of the spectral slope. Largest among these is the fact that, although the field as a whole has examined a range of tasks, to our knowledge, no single study has examined correlates of the spectral slope across a broad range of cognitive domains. This is important from the perspective of assessing the generality of spectral slope effects in aging, and the degree to which it may be more sensitive to declines in some functions than others (Dave et al., 2018, p. 40). In addition, prior studies that have examined cognitive correlates of the spectral slope have thus far focused on experimental tasks, as opposed to standardized measures, such as those used in neuropsychological assessment. Insofar as the spectral slope is of interest in terms of its translational potential as biomarker of age-related decline, it is beneficial to examine the degree to which variation in the slope relates to performance on tasks that have been validated for actual clinical decision-making. Thus, in light of these considerations, the primary goal of this study was to replicate and expand upon the prior findings relating flatter spectral slopes to cognitive performance declines in older adults, by using a larger battery of clinically validated cognitive tests. Following from the initial study by Voytek et al. (2015)–the only study to date which has demonstrated a mediating effect of the spectral slope– our primary hypothesis was that the slope of the EEG power spectrum (collected at rest), would mediate age-group differences in performance across diverse cognitive domains.

## Methods

### Participants

Forty-nine participants were recruited for the study: healthy young adults (YA; <35y; n=22); cognitively healthy older adults (OA; >59y; n=24); and older adults with mild cognitive impairment (MCI; n = 3) following approval of the University Institutional Review Board. We originally planned to recruit a comparable sample of older adults with amnestic MCI, which is considered a prodrome of Alzheimer’s disease (Levey et al., 2006). This sample would allow for comparison of neurological changes between neurologically healthy older adults and those with MCI. Due to the onset of COVID-19 during data collection, however, we had to postpone data collection in older adults with MCI as a safety precaution. As a result, the n=3 older adults with amnestic MCI are excluded from statistical analyses, and our focus is solely on our primary aim comparing cognitively intact younger and older adults in the current sample.

Following informed consent, participants completed a demographic survey, wherein they self-reported their education status, handedness, current medical status, and physical activity status. Older adults also self-reported their concurrent medications and sleep quality using The Pittsburgh Sleep Quality Index (PSQI), but these data we not obtained for younger adults.

Participants over 18 were recruited and did not have any impairments that would limit the function of their arms, hand, or legs. Participants were recruited from the local community by flyers and word of mouth recruitment. Older adults specifically were also recruited through a participant database maintained by the Center on Aging at the University of Utah. Participants with MCI were recruited from an ongoing study by one of the coauthors (KD), a licensed neuropsychologist, who also provided their diagnosis for amnestic MCI. Participants were excluded if they had any self-reported musculoskeletal, neurological, or perceptual impairments that they thought would affect their performance in these tasks, or any history of skin allergies (as a precaution for EEG data collection). In addition, participants were excluded if they had any medical history of severe cognitive impairment (e.g., dementia) or psychiatric conditions (e.g., severe depression, bipolar disorder, substance abuse).

### Tasks and Measures

Participants completed a battery of cognitive and motor tasks: the RBANS to assess cognition, a maximum grip-force task, a simulated feeding task (Schaefer et al., 2015; Schaefer & Hengge, 2016), and a sensorimotor test of standing balance. However, to address our primary aim the current study focuses only on the resting EEG data and the behavioral data from the RBANS.

### Cognitive Assessment

#### RBANS

This individually-administered, paper and pencil battery has been validated to assess cognitive function in adults ages 20-89 (Randolph et al., 1998). It is a reliable and valid assessment of cognitive status for detecting dementia (Duff et al., 2008), discriminating among individual differences in healthy older adults (England et al., 2014; Patton et al., 2003), and in similarly aged clinical populations (Larson et al., 2005). Furthermore, there is good evidence of convergent validity to show that RBANS scores are associated with instrumental activities of daily living (Freilich & Hyer, 2007; Larson et al., 2005). The RBANS consists of 12 subtests that load onto 5 indices of various cognitive domains: immediate memory, visuospatial/constructional ability, language, attention, and delayed memory. To ensure consistency of scoring, all tests were scored by the same author, AP. Although age-corrected normative data is available, we elected to use non-age-corrected raw scores so that we could compare age-groups on their absolute performance (rather than performance relative to age-matched peers).

### EEG Processing and Aggregation

Scalp EEG was collected from a 32-channel actiCAP active electrode system housed in a 64-channel cap and amplified with a BrainAmp DC amplifier (BrainProducts GmbH). The resting EEG data was collected for two minutes in each of two different conditions; eyes open and eyes closed. To reduce the potential for artifacts and make the parameterization of the power spectrum easier, we a priori chose to analyze only on data from eyes-closed rest. Participants were seated in each condition and instructed to, “Clear their minds and relax”. In the eyes open condition, all participants were oriented towards a fixation cross that was taped to a white wall approximately 2m away. The electrodes were labelled in accordance with the standard 10-10 geodesic montage (Oostenveld & Praamstra, 2001). The specific electrode sites included, Fp1, Fz, F3, F7, FT9, FC5, FC1, C3, T7, TP9, CP5, CP1, Pz, P3, P7, O1, Oz, O2, P4, P8, TP10, CP6, CP2, Cz, C4, T8, FT10, FC6, FC2, F4, F8, and Fp2. The data were online referenced to the right ear, with a common ground on the left ear. The electrode impedances were maintained below 25kΩ, with a sampling frequency of 1000Hz.

EEG data processing was conducted with the BrainVision Analyzer 2.1.2 software (BrainProducts GmbH). No online filters were applied, but offline the data were band-pass filtered between 0.1 and 40 Hz with 24-dB rolloffs with a 60 Hz notch filter. The data were manually inspected, and eyeblinks marked for ICA-based ocular correction within BrainVision Analyzer software (BrainProducts 2013). This function was run with the FP2 electrode serving as the VEOG and the HEOG electrode. Using Welch’s method, the data were epoched in 1-s segments, which overlapped by 50%. Following that, segments with artifacts were removed (average of 91% segments retained per person, SD = 6.73%). The remaining segments were subjected to a fast Fourier transformation using 0.977 Hz bins and a Hamming window with a 50% taper (Welch, 1967). The data were then truncated between 2-25 Hz to exclude gamma-band power and higher frequency muscle activation and were then averaged within sets of electrodes to get the average power spectrum in each region (Frontal: F7, F3, Fz, F4, F8; Central: C3, Cz, C4; Parietal: P7, P3, Pz, P4, P8; and Occipital: O1, Oz, O2).

### Calculation of the Exponent and Offsets

To compute the exponents and offsets for each participant, we used the spectral parameterization algorithm (*specparam* toolbox), which decomposes the power spectral density into the aperiodic background (exponent and offset) and the periodic peaks (Donoghue et al., 2020). The code for this algorithm is available as an open source *specparam* toolbox (https://fooof-tools.github.io/). The data were truncated to 2-25 Hz, and the *offsets* and *exponent* were extracted from the average power spectrum in each brain region for each participant. To ensure consistency of data processing, the EEG data was parametrized with the “*aperiodic_mode*” set at ‘fixed’, the width the peaks limited to 8, and the minimum peak height set to 0.05. The choice of parameters led to a median *r* ^*2*^= 0.96, IQR=[0.92,0.98] across all regions. Thus, although other processing parameters could have been chosen, we achieved good spectral parameterization across participants and regions.

### Statistical Analysis

All statistical analyses were conducted in R v4.1.0 (R Core Team, 2021; R Studio Team, 2020; Wickham et al., 2019). Following the extraction of the exponents and offsets, we used mixed-effects regression (Bates et al., 2014), with fixed effects of age-group and brain region, and their interactions. A random effect of Subject accounted for the within-subject nature of the Region factor.

To assess age-group differences in cognitive performance, we used independent samples t-tests to compare performance on the raw RBANS scores between age groups. *A priori*, we chose to conduct one-tailed tests as past data strongly suggest that older adults should generally score worse than younger adults. We tested the effect of age-group for RBANS domain-composite scores, calculated from the percentage of the maximum possible score on each subtest. Note that this is a departure from the way RBANS scores are normally analyzed, which requires age-normalizing the scores. Because our planned analyses require us to test the effect of age and then its (potential) mediation, we chose not to age-normalize the RBANS scores. Thus, we focus on the full RBANS composite score (percentile scores averaged across all domains) and domain composite scores (percentile scores combining subtests in each domain).

Finally, we used mediation analysis with non-parametric bootstrapping to assess the mediating effect of the exponent on age-related declines in cognition (Imai et al., 2010; Tingley et al., 2014). *A priori*, we chose to focus on the exponents from the frontal electrodes only, reflecting our interest in higher cognitive function. The exponents were extracted from the *specparam* toolbox (one frontal exponent value for each participant) and then used as a mediator in a series of regression models. Cognitive domain performance was regressed onto age-group, the exponent was regressed onto age group, and cognitive test performance was regressed onto age-group and the exponent. Using k=5,000 iterations, these models were bootstrapped to estimate the total effect, the average direct effect, and the average mediation effect.

To ensure reproducibility, de-identified EEG and behavioral data, plus all analysis code are available from the corresponding author’s GitHub account (https://github.com/keithlohse/spectral_slopes_aging).

## Results

### Demographics

Forty-nine participants were recruited from within the university and the larger community. However, due to a data storage issue, we lost EEG data for one YA participant, bringing our total YA n=21, and one OA participant, bringing our total OA n=23. Demographic statistics for these participants are shown in Table 1. Data from the n=3 older adults with MCI were too limited to include in statistical analyses. All subsequent analyses are based on the N=44 cognitively intact older and younger adults.

**Table 1.** Summary statistics for the participant demographic surveys.

| Variable | Younger Adults | Older Adults |
| --- | --- | --- |
| Participants (n) | 21 | 23 |
| Age, mean (SD) | 23.29(3.47) | 70.83(5.77) |
| Age, range | 19, 33 | 59, 83 |
| Male (n) | 11 | 4 |
| Education level (years), mean (SD) | 14.93 (1.58) | 15.45(2.61) |
| # of Current Medications | *Not Collected* | 1 to 23 |
| No. of medications, mean (SD) | *Not Collected* | 5.55(5.26) |
| Average Maximum Grip Force (kgf) | 20.5 (6.5) | 12.4 (4.2) |
| Self-Reported Physical Activity |  |  |
| Vigorous PA (Days per week) | 2.98(1.75) | 2.22(2.33) |
| Vigorous PA (Hours per day) | 1.21(0.64) | 0.87(0.91) |
| Moderate PA (Days per week) | 3.67(1.72) | 4.15(2.52) |
| Moderate PA (Hours per day) | 1.42(0.83) | 1.51(1.22) |
| Light PA (Days per week) | 5.76(2.12) | 5.43(2.42) |
| Light PA (Hours per day) | 3.74(3.15) | 2.91(2.08) |
| Resistance training (Days per week) | 2.52(2.09) | 1.17(1.59) |
| Sleep Quality, mean (SD) | *Not Collected* | 5.33(3.57) |

Note that details of medications taken and the Pittsburgh Sleep Quality Index were only added to the study protocol after data collection in the younger adults was finished. As such, these data are only available for the older adults.

### Age-Related Differences in EEG Power Spectra

To measure age-related changes in the exponent in different brain regions, a linear mixed-effect regression model with fixed effects of age-group and brain region, and their interactions revealed a significant main effect of Age-Group (F(1,44) = 12.56, *p* < 0.001) and brain Region (F(3,132) = 8.70, *p* < 0.001). These effects were superseded by the Age-Group x Region interaction (F(3,132) = 10.09, *p* < 0.001). To decompose this interaction, we subset the data into regions, and performed linear regressions with fixed-effects of Age-Group.

In the frontal region, there was a significant main effect of Age-Group, F(1,42)=13.28, p <0.001. The exponents in the frontal region were lower in older adults (i.e., flatter power spectra) as compared to the younger adults, see Figure 2B.

**Figure 2.**
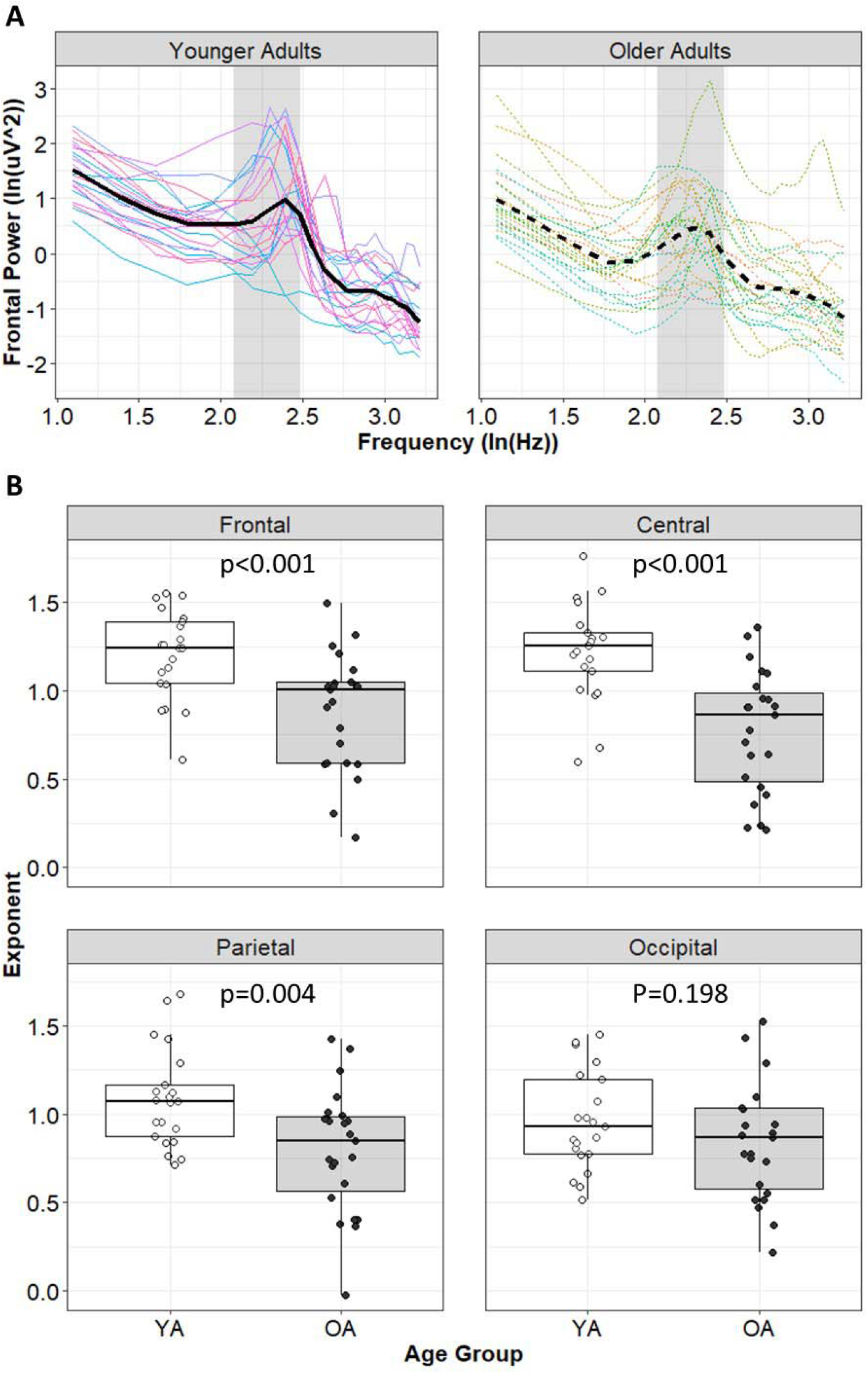
(A) Average power spectra across the frontal electrodes (F7, F3, Fz, F4, F8) as function of age-group, shown in log-log space. Thin colored lines show individuals, thicker black lines show the average power spectra for the whole group (line dashing varies with group). (B) Estimated exponents from each region as a function of age group. P-values are based on a linear model regressing exponents onto a contrast coded age group variable in each region. YA = younger adults; OA = older adults. The canonical alpha band (8-12 Hz) is highlighted in Panel A.

In the central region, there was a there was a significant main effect of Age-Group, F(1,42)=22.00, p <0.001. The exponents in the central region were reliably lower in older adults as compared to the younger adults, see Figure 2B.

In the parietal region, there was a there was a significant main effect of Age-Group, F(1,42)=9.24, p = 0.004. The exponents in the parietal region were reliably lower in older adults as compared to the younger adults. In the occipital region, however, no significant effect of Age-Group was observed, F(1,42) = 1.71, p = 0.198. Both results are shown in Figure 2B

### Age-Related Differences in Cognition

As shown in Figure 3, there was considerable variability both within and between groups on the different scales of the RBANS. For the RBANS composite score, there was a statistically significant difference between younger and older adults (t(41.6) = −1.87, p=0.034), with older adults performing worse (d=-0.40). Delving into the specific cognitive domains of the RBANS, only Attention (t(38.1) = − 2.86, p = 0.003, d=-0.61) and Delayed Memory (t(41.8) = −1.84, p=0.036, d=-0.39) showed statistically significant differences, whereas differences for Immediate Memory (t(40.3)=-0.38, p = 0.353, d=-0.08), Visuospatial/Constructional ability (t(42.0)=-1.01, p = 0.158, d=-0.21), and Language (t(39.7) = 0.62, p = 0.730, d=0.13) were not statistically significant.

**Figure 3.**
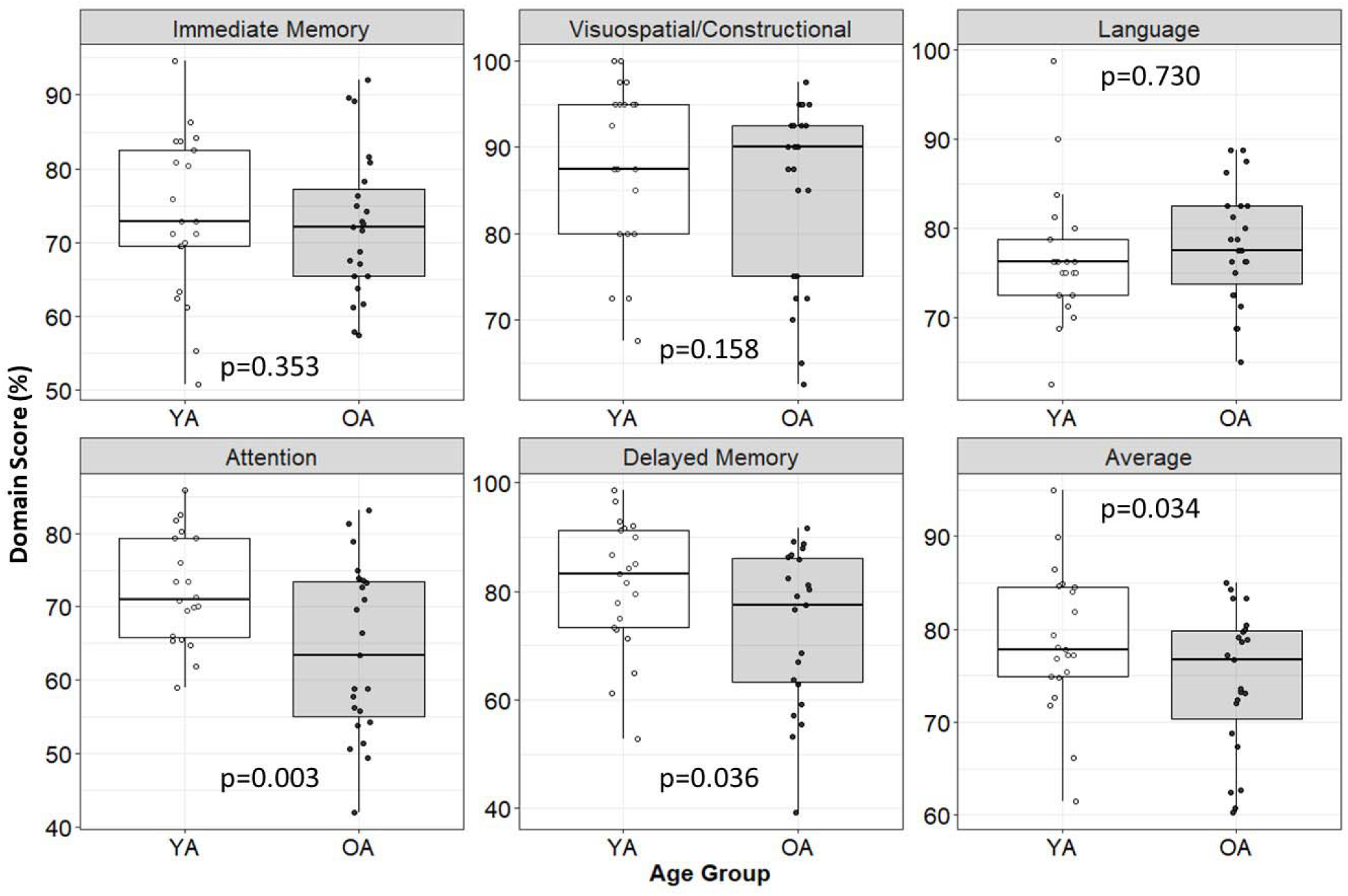
RBANS scores for each cognitive domain as a function of group: Immediate memory, Visuospatial/ Constructional abilities, Language, Attention, and Delayed Memory. P-values are based on directional Welch’s Two Sample t-test (not assuming equal variances), where *H*_0_: *OA* > *YA. YA* = younger adults; OA = older adults.

### Does the spectral slope mediate age-related changes in cognition?

Given our *a priori* predictions, exponents from the frontal region were tested as potential mediators for age-group differences in the RBANS. As shown in Table 2, we tested only those RBANS scales that showed statistically significant differences between age-groups. Contrary to our predictions, neither total RBANS scores (p=0.170), the Delayed Memory domain (p=0.183), nor the Attention domain (p=0.259) showed a statistically significant mediation effect. See Figure 4A-C.

**Table 2.** Testing the exponent of the power spectrum as a mediator of age-related differences using non-parametric bootstrapping to estimate mediation effects.

| **Dependent Variable** | **Total Effect** |  | **Direct Effect** |  | **Average Mediation Effect** |  |
| --- | --- | --- | --- | --- | --- | --- |
|  | **Estimate** | **p-value** | **Estimate** | **p-value** | **Estimate** | **p-value** |
| Average (%) | 4.34 | 0.050 | 2.50 | 0.340 | 1.84 | 0.170 |
| Delayed Memory (%) | 7.29 | 0.057 | 4.04 | 0.378 | 3.25 | 0.183 |
| Attention (%) | 8.36 | 0.003 | 6.29 | 0.070 | 2.06 | 0.259 |
| -- Coding Subtest (%) | 17.32 | <0.001 | 13.92 | <0.001 | 3.40 | 0.034 |
| -- Digit Span Subtest (%) | -0.61 | 0.860 | -1.34 | 0.800 | 0.73 | 0.800 |

\*Note that the total effect reflects the effect of both the grouping variable (Age Group) and the mediator (spectral power Exponent) on the outcome (domains of cognition). The direct effect is the effect of the group variable with the mediator removed, and the mediation effect is a statistical test of the exponent as a mediator.

**Figure 4.**
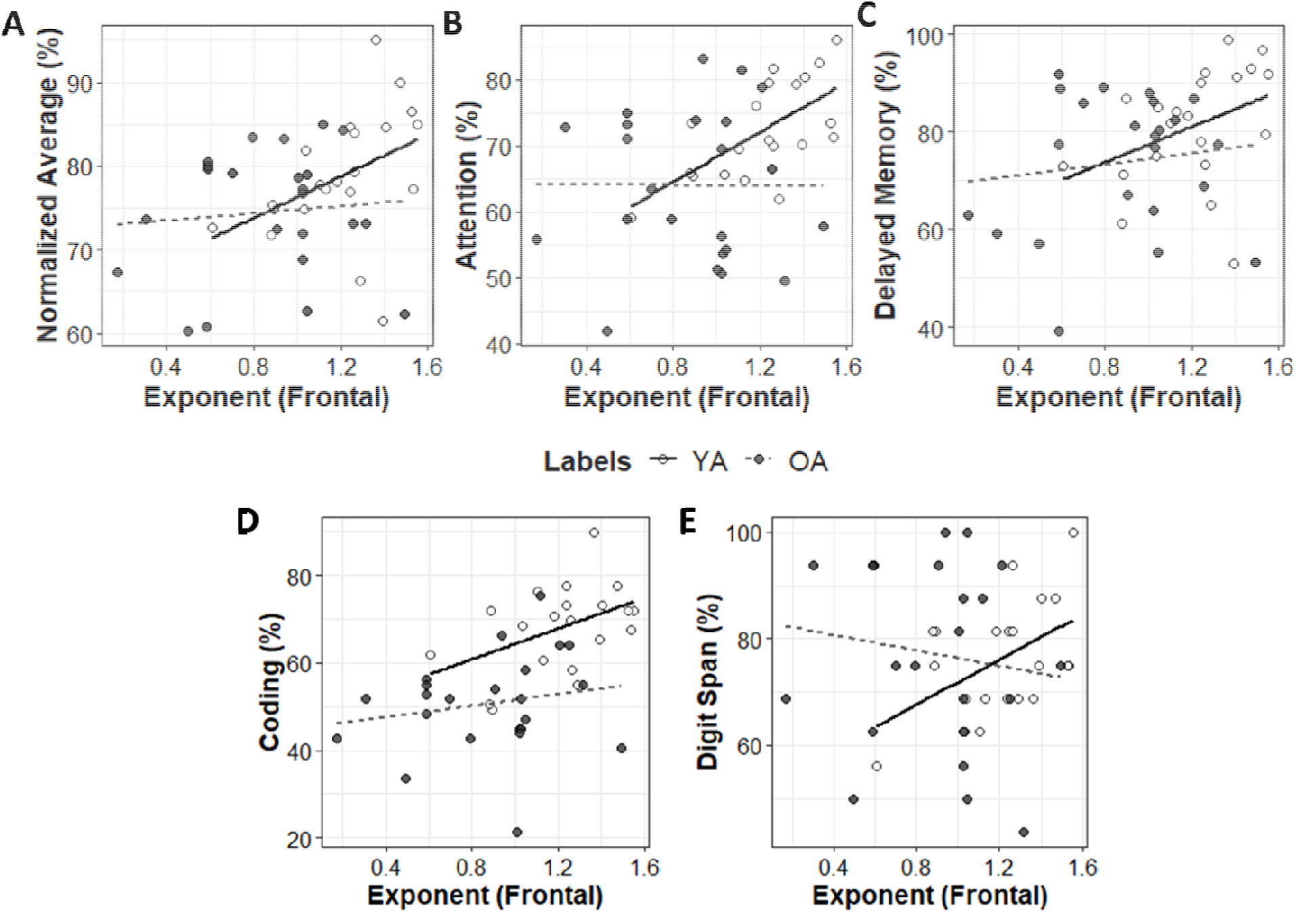
RBANS total (A), Attention domain (B), Delayed Memory domain (C), Coding subtest (D), and Digit Span subtest (E) scores as a function of the spectral slope across the frontal electrodes. These subtests were selected for the statistically significant differences between younger (YA) and older adults (OA) shown in Figure 3. Lines indicate the ordinary least squares regression line within each group.

In an earlier version of these analyses published in a pre-print (Pathania, Clark, et al., 2021), we did find evidence of statistically significant mediations. However, those analyses used regression-based methods to estimate the slope of power spectrum (rather than spectral parameterization) and analyzed the RBANS at the level of specific subtests rather than scales. To better understand these discrepant results, we focused on the Attention scale, which showed the largest age-related differences, and ran mediation analyses on its constituent subtests: the Coding test and the Digit Span test (see Figure 4D and E). As shown in Table 2, there was evidence of mediation for the Coding subtest (p=0.034), but not for the Digit Span subtest (p=0.800). Thus, across our pre-print and the current analysis, there was evidence that the exponent of the frontal power spectrum mediated age-related differences in the coding subscale. This suggests that the mediation is not an artifact of the method of estimation (regression in the pre-print, spectral parameterization here) and is instead something unique about the Coding subtest that is not shared with the Digit Span.

## Discussion

Consistent with the neural-noise hypothesis of aging, our data showed that (1) the slope of the resting EEG power spectrum is flatter in older compared to younger adults (replicating past work) and (2) individual differences in the slope of the EEG power spectrum mediated age-related differences in a clinically validated assessment of information processing speed (extending past work). However, it is important to note that this pattern was not evident across the five different domains of the RBANS. Contrary to our predictions, none of the RBANS domains showed a reliable mediation effect. Since the Attention domain showed the largest age-related differences, we performed mediation analysis with its constituent subscales, Coding and Digit Span. The effect was apparent only in the Coding subtest. Other domains/subtests of the RBANS were not examined, as we did not find reliable evidence of cognitive differences between the performance of younger adults and older adults. Thus, combining the current data with past work, it appears that the flattening of the exponent with age is a reliable effect (Tran et al., 2020; Voytek et al., 2015; Waschke et al., 2017). Further, the exponent is related to cognition not only in tightly controlled laboratory tasks (Thuwal et al., 2021, Tran et al., 2020; Voytek et al., 2015), but also in validated clinical tests (the Coding subtest of the RBANS in the present data).

The obtained pattern of results raises important questions about how the findings pertaining to Coding subtest of the RBANS should be interpreted. The Coding subtest falls under the “Attention” domain. Interestingly, the Digit Span is the other subtest in the Attention domain, and we found no evidence of a mediation effect with respect to Digit Span scores. Ultimately, the reason for the strong association with the Coding subtest is not clear, but we posit a few possible (non-exclusive) explanations. First, the Coding subtest has the strictest time requirements of any subtest on the RBANS, and is directly analogous to other symbol substitution tasks that are commonly used to assess processing speed in neuropsychology (Larrabee, 2017). To perform the task efficiently, participants need to memorize the correspondence between symbols presented on the paper and the numbers those symbols represent (i.e., a symbol-number pairing). As the task goes on, more efficient participants are able to encode the symbol-number pairings into working memory, as opposed to repeatedly alternating between the symbol key and the response field. Thus, these tasks require the rapid integration of various sub-processes, including visual scanning, working memory, sustained attention, and visuomotor coordination, and consequently quantify an individual’s ability to rapidly learn, coordinate, and execute a set of simpler sub-tasks (Lezak et al., 2004).

A second, perhaps complementary, perspective on these measures highlights the importance of executive functions, via cognitive control and working memory, in allowing individuals to rapidly coordinate the necessary sub-processes when faced with a novel task and timed testing conditions (Koziol & Budding, 2009). Both of these perspectives are consistent with the large literatures that highlight: (1) the relative prominence of changes in the (interrelated) functions of processing speed, working memory, and fluid cognition with greater age (Salthouse & Davis, 2006), and (2) the effects of clinical conditions affecting distributed processing and/or white matter integrity on timed symbol-substitution tasks (e.g., DeLuca et al., 2004; Kinnunen et al., 2011; Lezak et al., 2004).

Interestingly, we did not find evidence for the exponent mediating age-related differences in delayed memory. From these pilot data, we cannot conclude that there is no relationship between exponents and delayed memory, but we found no evidence for a mediating effect in the current sample. There are several reasons why there might be no evidence of mediation in delayed memory despite a relatively large age-related difference. It could be that there truly is no mediating effect in delayed memory in the population or it could be that there is an effect but we lacked the power to detect it. This second option is an important consideration because in the present sample the Attention domain showed the largest age-related differences. If differences in other domains were larger (i.e., between group variance increased or within group variation reduced) that would increase the power to detect a mediation if one exists in that domain. At present, however, we only find evidence of a mediation effect for the Coding subtest, which leans most heavily on speeded information processing.

Finally, it is unfortunate that we were unable to collect data from our sample of older adults with MCI. The descriptive data for three participants are interesting, but far too limited to allow a substantial qualitative interpretation. Based on the relationship between resting EEG exponents and performance in cognitively intact older adults, however, we do think that these pilot data warrant further research into the clinical utility of the resting spectral slope as a biomarker of cognitive decline. We have shown that resting EEG exponents are sensitive to individual differences in some aspects of cognition. Combined with past work showing how frequency-based EEG measures can longitudinally predict the conversion from MCI to Alzheimer’s disease (Engedal et al., 2020; Poil et al., 2013), we think the present data warrant longitudinal studies to see if the EEG exponents possibly outperform other biomarkers, or if the predictive utility of other biomarkers is improved after accounting for the EEG exponent/offset (Donoghue et al., 2020).

## Limitations

There are a number of limitations in the present study of which we need to be cognizant. First and foremost, we have a limited sample size and we need to careful in the interpretation of our hypothesis test results. Due to relatively low power in some analyses, significant results may not be representative of the real underlying effects, and non-significant results may have simply lacked adequate statistical power (Button et al., 2013; Lohse et al., 2016). Finally, although the RBANS does tap multiple cognitive domains, it does not have pure measures of executive functioning. Given the present results, future studies might examine if executive function specifically is influenced by age and spectral slope.

## Conclusions

Consistent with the neural-noise hypothesis of aging, we replicated earlier work showing that the resting EEG exponents become flatter with age and that individual differences in the exponents explain individual differences in cognition. We also extend this past work by demonstrating these effects in clinically validated assessments of cognitive status. However, the mediation effect was only present in a test of information processing speed, suggesting that these effects are not broadly applicable to all areas of cognition.

## Data Availability

All data and code are shared at https://github.com/keithlohse/spectral_slopes_aging

